# A connection-level genetic atlas of human cortical functional connectivity at rest and during a task

**DOI:** 10.64898/2026.10.28.26366764

**Authors:** Shushan Wu, Bingxin Zhao, Zhiwen Jiang, Akemi Hara, Zehui Sun, Xiaoqi Li, Zhaoyang Zhang, Xiaoyu Qi, Isaac Pan, Yuqi Cheng, Yukang Jiang, Borui Tang, Shuai Huang, Shan Gao, Shucheng Liu, Shilin Yu, Yuhao Zhou, Peter Guan, Mingcheng Hu, Yuyang Deng, Yun Li, Jason Stein, Patrick F Sullivan, Tengfei Li, Hongtu Zhu

## Abstract

Genetic studies of cortical functional connectivity have analyzed aggregated traits, which fails to show which connections a variant acts on. We present a connection-level genetic atlas: genome-wide association analyses of 64,620 connections among 360 cortical areas in UK Biobank participants of European ancestry at rest (*n* = 52,983) and during an emotion-processing task (*n* = 45,010), with replication in an independent sample (*n* = 15,546). We identified 247 resting and 96 task loci; 193 resting loci were new to fMRI genetics and 112 replicated. Most loci affected connections between networks. Colocalization linked *APOE* ε4 to lowered posterior default mode connectivity and a cognition locus to left posterior temporal (PHT) connections; the right PHT was a replicated hub. Genes near the loci included 155 drug targets, 50 with psychiatric indications and 15 targeted by approved psychiatric drugs. Resting connectivity was more heritable and colocalized more often with complex traits than task connectivity, whereas task connectivity was more strongly associated with circulating metabolites. The atlas provides summary statistics for every connection in two brain states.

---

The cerebral cortex is organized into areas and networks with distinct functions^1,2^. Functional magnetic resonance imaging (fMRI) measures how these areas interact as functional connectivity, the correlation of blood-oxygen-level-dependent (BOLD) signals between areas^3,4^. Resting-state fMRI measures this coupling without a task, and task fMRI measures it while a task is performed. Connectivity is altered in schizophrenia^5,6^, Alzheimer’s disease^7,8^, autism spectrum disorder^9,10^, and major depressive disorder^11,12^. It is also moderately heritable^13–15^ but less reliable than structural imaging measures^16,17^. Genetic studies of connectivity therefore need biobank-scale samples, such as the UK Biobank (UKB)^18,19^.

Earlier studies have mapped genetic loci for large-scale connectivity. Genome-wide association studies (GWAS) in UKB for resting fMRI found loci near genes such as *EPHA3* and *PAX8* and colocalized the *APOE* ε4 locus with Alzheimer’s disease^20–22^. These studies analyzed components and networks, which combine many connections. Components derived by independent component analysis (ICA) span several areas and networks, so a genetic signal is hard to attribute to a specific region^22,23^. Parcellation-based studies analyzed a few networks or network-level traits^24,25^, and a recent study of individual connections used a coarser parcellation of 82 regions^26^. Combining connections dilutes the effect of a variant that acts on only some of them, but testing every connection multiplies the computational and multiple-testing burden. Three questions therefore remain open: which connections genetic variants act on, whether loci for cognition and disease act on the connections that carry the polygenic signal of those traits, and whether the answers differ between rest and task.

Here we characterize the genetic architecture of individual cortical connections at rest and during a task. We performed GWAS of all 64,620 functional connections (edges) among the 360 areas of the Glasser atlas^1,27^ in resting (*n* = 52,983) and task fMRI (*n* = 45,010) and tested replicated of the resting loci in UKB imaging phase 7 (an independent sample with similar methodology, *n* = 15,546). A low-dimensional representation (LDR) framework based on principal component analysis made this feasible: it tests a compact set of components and reconstructs the effect of each variant on every edge^28^.

Significance is calibrated by the effective number of independent edges^28,29^. We call the edges associated with a locus its footprint. Footprints show which connections each locus acts on. To test whether loci for cognition and disease act on the connections that carry the polygenic signal of their traits, we compare footprints with edge-level genetic correlation maps of 36 traits, a test we call circuit closure. To test whether genetic effects differ between rest and task, we compare the two states edge by edge. We then link the loci to genes with colocalization, molecular quantitative trait loci (QTLs), and Mendelian randomization, and to candidate drug targets with drug-target databases. The atlas provides summary statistics for every connection in both brain states.

## RESULTS

### Overview of imaging traits and analyses

The 360 areas of the Glasser atlas^1^ were grouped into 12 functional networks^30^ (**Fig. 1A** and **Table S1**). In each state, the edges were divided into four parts. Within each part, the top 300 principal components (LDRs) were tested for association, and the effect of each variant on every edge was reconstructed from them. The discovery samples comprised UKB participants of European ancestry (Methods) with resting (*n* = 52,983) or task fMRI (*n* = 45,010). Imaging phase 7 provided an independent replication sample of 15,546 participants (**Fig. 1A**). Study-wide significance was *P* < 5 × 10^-8^/*M*_eff_, where *M*_eff_ is the effective number of independent edges, computed from all eigenvalues of each part (**Methods**). *M*_eff_ was 46.7 at rest and 26.9 during the task, which gave thresholds of *P* < 1.07 × 10^-9^ and 1.86 × 10^-9^. We call the edges associated with a locus its footprint (**Fig. 1B**). The downstream analyses are outlined in **Fig. 1C–E**.

**Fig. 1.**
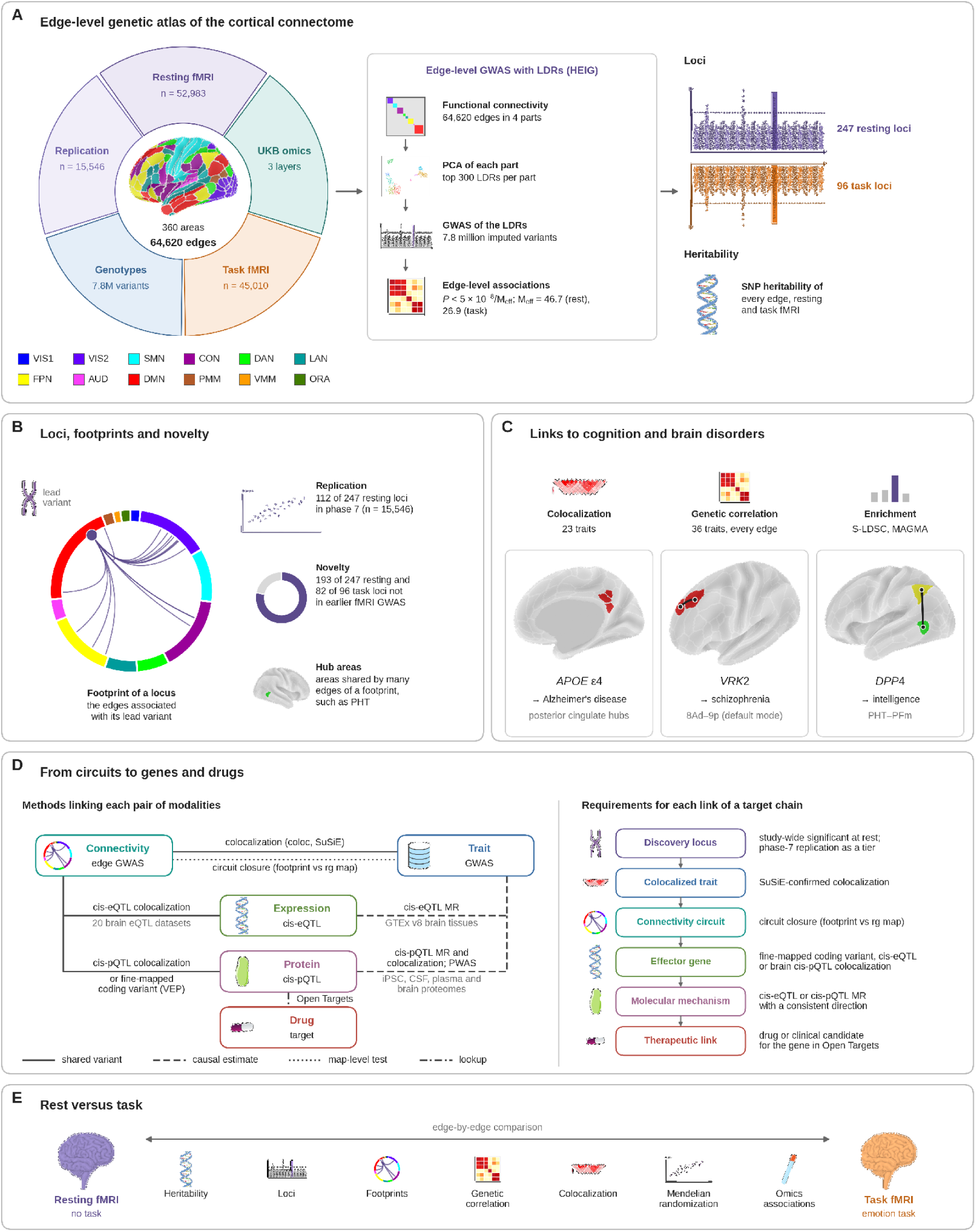
Study design and analytical framework. **(A)** Edge-level genetic atlas of the cortical connectome. The ring shows the UK Biobank (UKB) data: resting fMRI from imaging phases 1 to 6 (*n* = 52,983) and task fMRI (*n* = 45,010) for discovery, resting fMRI from imaging phase 7 for replication (*n* = 15,546), 7.8 million imputed variants, and three omics layers. The 360 areas of the Glasser360 atlas^1^, colored by their 12 functional networks^30^, form 64,620 edges. In each state, genome-wide association of low-dimensional representations (LDRs), from principal component analysis of four parts of 16,155 edges in HEIG, gave the association of each variant with every edge (study-wide *P* < 5 × 10^-8^/*M*_eff_, with *M*_eff_ the effective number of independent edges; Figs. 2 and 3). **(B)** Loci, footprints, and novelty (Fig. 3 and Tables S2 and S3). **(C)** Links to cognition and brain disorders (Figs. 4–6). The maps show the posterior cingulate hubs of the *APOE* ε4 footprint and the left 8Ad–9p and PHT–PFm connections at the *VRK2* and *DPP4* loci. **(D)** From circuits to genes and drugs (Fig. 7 and Table 1). Left, the evidence that links each pair of levels through the lead variant, with line style marking its type; right, the requirement for each link of a target chain. **(E)** Rest versus task: resting-state and task-state connectivity compared edge by edge (Fig. 8 and Figs. S1 and S2). Details are in the Supplementary Note.

**Table 1.**
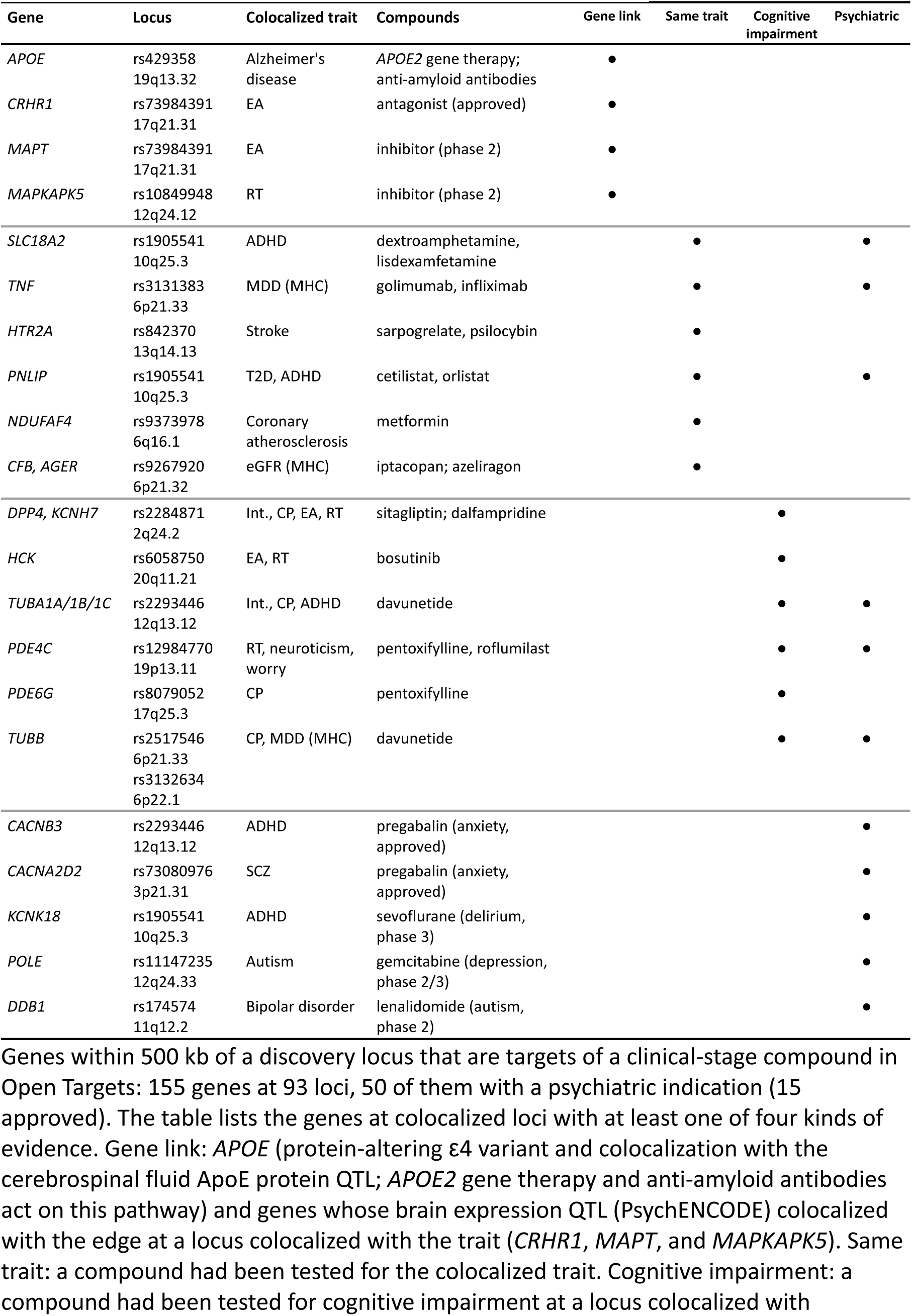
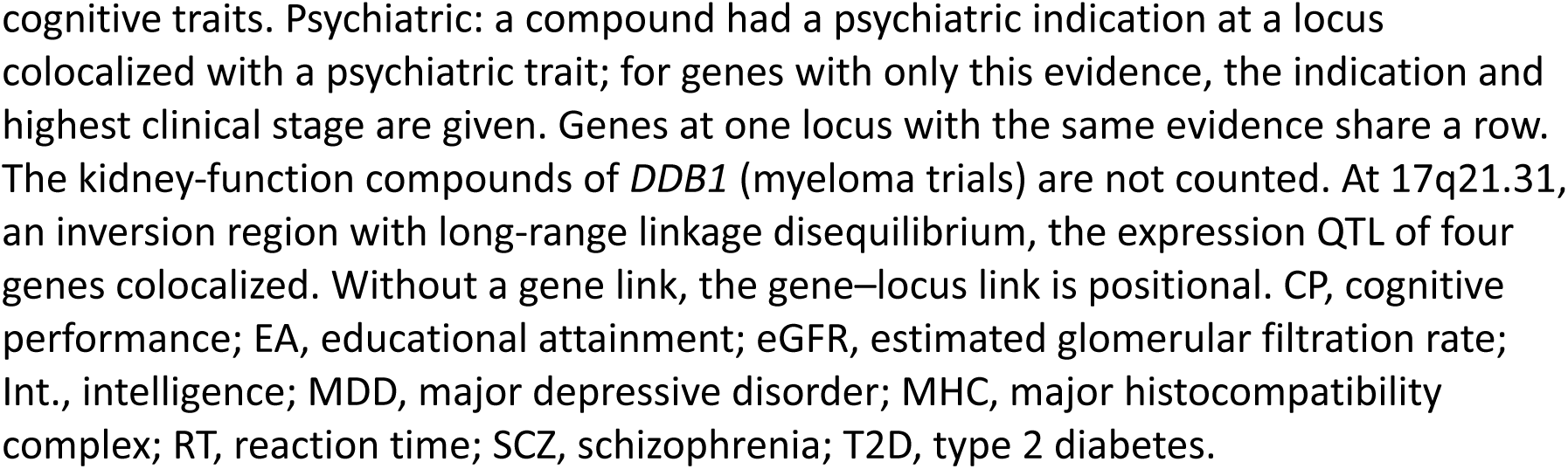
Drug targets near the resting-state loci.

### Heritability of cortical functional connectivity at rest and during a task

We first estimated the SNP-heritability of each edge from the LDR summary statistics and tested it at a false discovery rate (FDR) of 5% across edges. Mean heritability was 8.9% at rest and 7.1% during the task, and heritability was significant for 94.8% and 99.0% of the edges with estimates (**Fig. 2A, B**). Over the significantly heritable edges, the mean resting heritability (9.3%) matched that of ICA-derived traits^22^. At rest, connections between networks were nearly as heritable as connections within networks (means over all edges with estimates, 8.7% versus 10.0%). The coupling between networks, which network-level traits average out, is therefore itself heritable. Resting heritability peaked in the posterior multimodal, dorsal attention, default mode, and frontoparietal networks (means over significant edges, 9.7% to 11.0%; **Fig. 2C**), whereas task heritability was more uniform across networks (5.7% to 7.9%; **Fig. 2D**). The default mode and frontoparietal networks belong to the triple network, which was also the most heritable among ICA-derived traits^22^. The most heritable areas were the left PGi, the PHT area of both hemispheres (**Fig. 2E**), the left TE1p, and the right STSvp, all in the inferior parietal and lateral temporal cortex. Connections of these areas offer the most power for genetic studies.

**Fig. 2.**
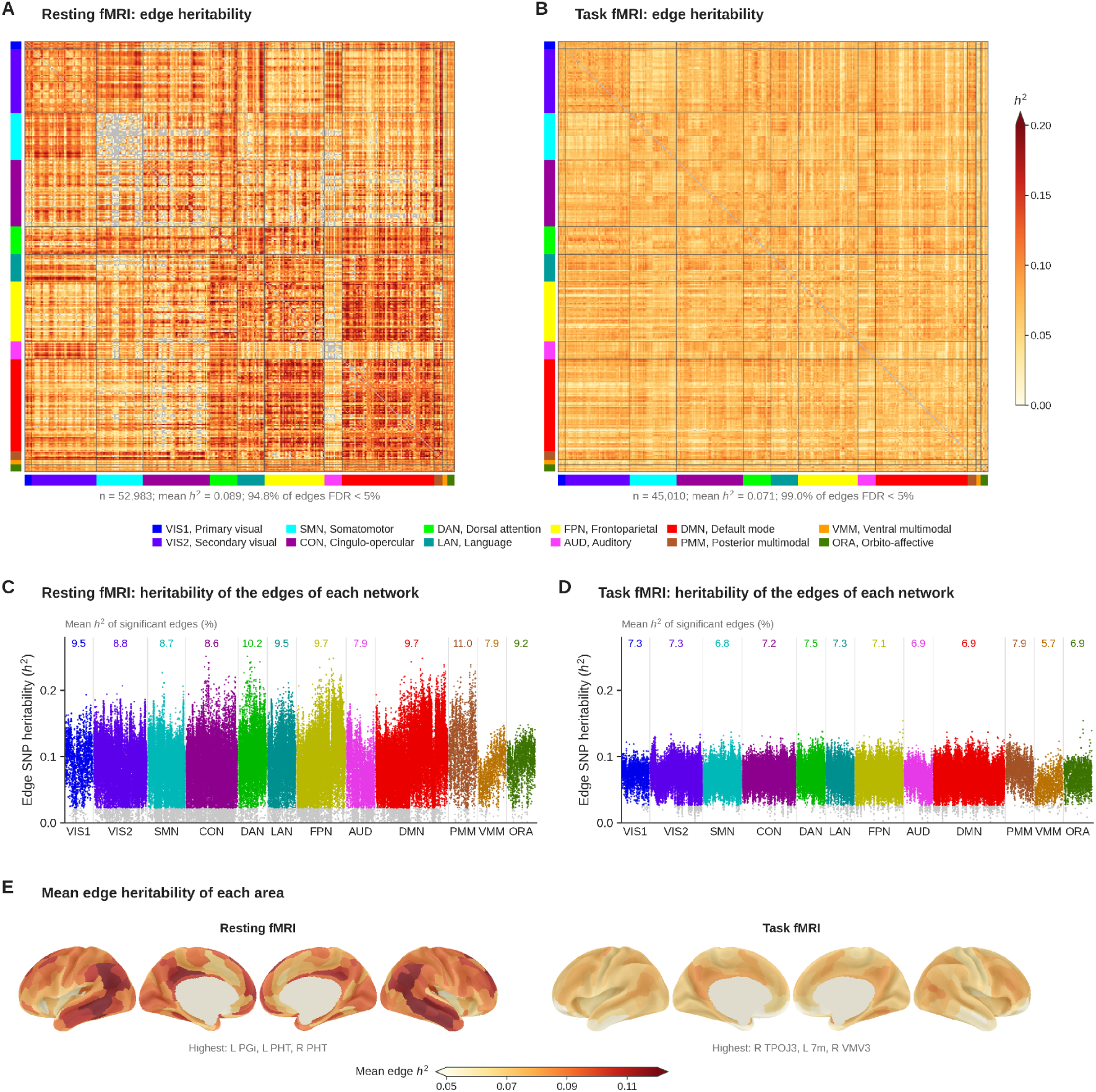
SNP heritability of edge-level functional connectivity in resting and task fMRI. **(A, B)** SNP heritability (*h*^2^) of the 64,620 edges in resting fMRI (**A**; *n* = 52,983) and task fMRI (**B**; *n* = 45,010), shown as 360 × 360 matrices in which the areas are ordered by network (colored bars). Edges without an estimate within the valid range and the diagonal are gray. The mean heritability and the percentage of the edges with estimates that had significant heritability (FDR < 5%) are given below each matrix. Values above 0.20 are shown in the darkest color. **(C, D)** Heritability of the edges of each network in resting (**C**) and task (**D**) fMRI. Each block contains all edges of one network, so an edge between two networks appears in both blocks. Colored points are edges with significant heritability (FDR < 5%), gray points are nonsignificant edges, and the numbers above the blocks give the mean heritability (%) of the significant edges. **(E)** Mean heritability of the edges of each area (over the edges with estimates) in resting and task fMRI, shown on the inflated cortical surface (left lateral, left medial, right medial, and right lateral views), on a color scale shared by the two states and clipped at 0.05 and 0.12. The three most heritable areas are listed below each map. Details are in the Supplementary Note.

### Genetic loci associated with edge-level functional connectivity

To find the variants behind this heritability, we selected variants that reached study-wide significance and were associated with more edges than expected by chance, and clumped them into independent loci (Methods). We identified 247 loci for resting fMRI and 96 for task fMRI (**Fig. 3A** and **Table S2**). Most loci affected few edges: footprints ranged from 1 to 24,888 edges (median 5) at rest and from 1 to 924 edges (median 2) during the task. For 176 resting (71%) and 85 task loci (89%), the strongest association was with a connection between network pairs (**Fig. 3B**). Such connections make up 87% of all edges, however, network-level traits capture them poorly. In both states, the strongest locus lay near *EPHA3* (3p11.1; **Fig. 3A, C**), which encodes an ephrin receptor involved in axon guidance^31^. This locus had been reported in earlier fMRI GWAS^21,22^. Its resting lead variant (rs35124509, *P* = 2.0 × 10^-89^) affected 2,889 edges, with hubs in the PHT area of both hemispheres and the left PGp. Its task lead variant affected 279 edges (**Fig. 3D**).

**Fig. 3.**
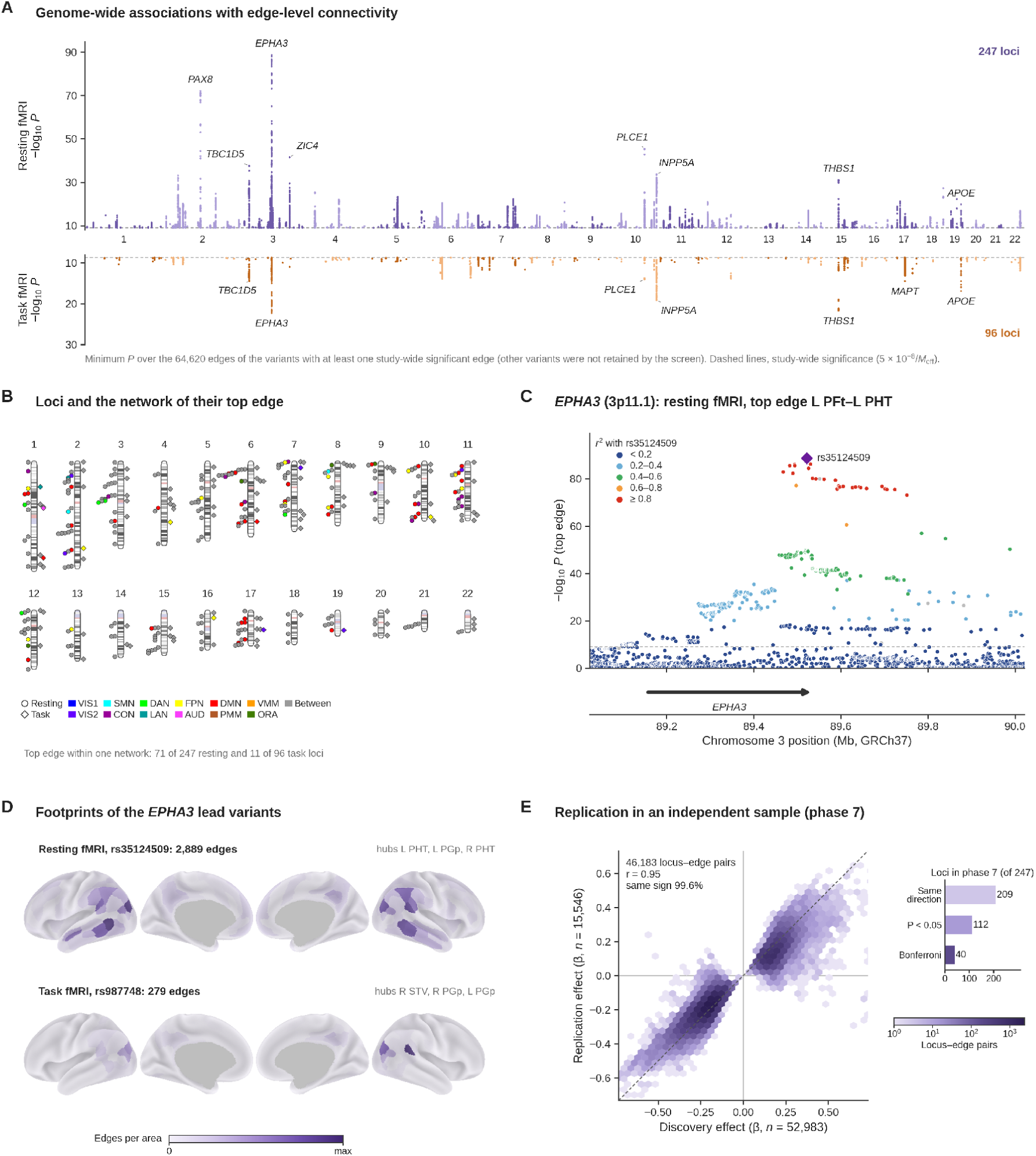
Genetic loci associated with edge-level functional connectivity. **(A)** Miami plot for the 64,620 edges in resting (top; *n* = 52,983) and task (bottom; *n* = 45,010) fMRI. For each variant with at least one study-wide significant edge, the smallest *P* value across edges is shown. Dashed lines, study-wide significance (1.07 × 10^-9^ for resting and 1.86 × 10^-9^ for task fMRI). Selected loci are labeled with a nearby protein-coding gene. **(B)** Chromosomal locations of the 247 resting (circles) and 96 task (diamonds) loci, colored by the network of the edge with the smallest *P* value of the lead variant across all edges when both areas belong to the same network and gray otherwise. Network abbreviations are as in Fig. 2. **(C)** Regional association plot of the 3p11.1 (*EPHA3*) locus, the most significant in both states, at the top resting edge of rs35124509 (left PFt–PHT; *P* = 2.0 × 10^-89^). Colors give the linkage disequilibrium (*r*^2^) with rs35124509 in the 1000 Genomes European panel; dashed line, study-wide significance. **(D)** Footprints of the *EPHA3* lead variants, rs35124509 at rest (2,889 study-wide significant edges) and rs987748 during the task (279 edges), shown as significant edges per area (each map scaled to its own maximum; top three areas listed). **(E)** Replication in UKB imaging phase 7 (*n* = 15,546). Hexagonal bins show the effects of the 247 lead variants at their 46,183 study-wide significant edges in the discovery and replication samples (Pearson *r* = 0.95; dashed line, equal effects). Inset, number of loci whose lead variant had the same direction of effect at its top edge within its discovery part (209), and also *P* < 0.05 (112) or *P* < 0.05/247 (40). Details are in the Supplementary Note.

In the phase 7 sample, we tested each resting lead variant at its top discovery edge. A locus replicated when the effect had the same direction and *P* < 0.05 (nominal level) or *P* < 0.05/247 (Bonferroni level). Of the 247 resting loci, 209 had the same direction of effect, 112 replicated at the nominal level, and 40 replicated at the Bonferroni level (**Fig. 3E**). The replication sample was 29% the size of the discovery sample. Replication extended beyond the top edges: across the 46,183 locus-edge pairs that were study-wide significant in the discovery sample, 99.6% of effects had the same sign in the replication sample, and effect sizes were correlated (*r* = 0.95). All 247 loci entered the downstream analyses (Supplementary Note), and **Table S2** gives the replication status of each locus.

The resting loci recovered most earlier findings: they lay within 500 kb of 41 of the 45 regions reported for ICA-derived resting traits^22^ and of all 19 regions with Bonferroni-significant fMRI associations in the BIG40 resource^21^. Most loci were new to fMRI genetics: they lay more than 500 kb from any study-wide significant fMRI association reported by earlier GWAS (**Table S3** and Supplementary Note). The new loci comprised 193 of the 247 resting loci, including 68 of the 112 replicated loci, and 82 of the 96 task loci. Of the 193 new resting loci, 149 had been reported for other brain-imaging traits, such as structural and diffusion measures, and 44 were new to brain imaging.

Stratified LD score regression placed the heritability of the 228 resting locus edges in active regulatory elements (H3K27ac peaks) of cortical neurons and glia (2.85-fold enrichment, enrichment *P* = 1.5 × 10^-9^; **Table S4**)^32,33^. The enrichment was similar in astrocytes, oligodendrocytes, and neurons (3.5- to 4.9-fold), in line with the enrichment of ICA-derived traits in fetal brain regulatory regions^22^. Gene-set analysis^34^ implicated neural development through genes near the lead variants (17 gene sets at an FDR of 5%; **Table S5** and Supplementary Note).

### Shared genetic influences with brain disorders and cognition

To link the loci to brain disorders and cognition, we tested each locus for a causal variant shared with 23 complex traits and diseases by Bayesian colocalization at the top edge of its lead variant. Strong colocalization required a posterior probability of a shared variant (PPH4) of at least 0.8 that exceeded that of distinct variants (PPH3)^35^. Across the 247 resting loci, 135 locus-trait pairs at 66 loci showed strong colocalization (**Table S6**). SuSiE-based colocalization^36^, which allows several causal variants, confirmed 76 pairs at 44 loci, 28 of them new to fMRI GWAS.

The strongest signal shared with a brain disorder was rs429358 (19q13.32), which with rs7412 defines the *APOE* ε4 allele, the major common risk factor for late-onset Alzheimer’s disease^37,38^. In both states, the ε4 association with connectivity colocalized with Alzheimer’s disease and related dementias^39^, with rs429358 as the shared variant (**Fig. 4A, B**). The resting association replicated in phase 7 (*P* = 6.3 × 10^-6^). An ICA-based study had also linked this locus to Alzheimer’s disease^22^; the atlas shows which connections it affects and in which direction. The ε4 allele lowered connectivity at all 589 significant resting edges and all 149 task edges (**Fig. 4C, D**). At rest, these edges lay in the default mode (333) and secondary visual (236) networks, with hubs in the right posterior cingulate areas 31pd, v23ab, and 31pv. During the task, they lay in the secondary visual network (**Fig. 4C–E**). Across all edges, the effects were negative at 79.6% of resting and 62.8% of task edges (**Fig. 4F**). The ε4 allele was also weakly associated with mean connectivity (*z* = −2.30). Because most participants were free of dementia, the lower posterior default mode and visual connectivity of ε4 carriers may mark genetic risk before clinical disease. Whether these connections carry the genetic correlation of Alzheimer’s disease is tested below with circuit closure.

**Fig. 4.**
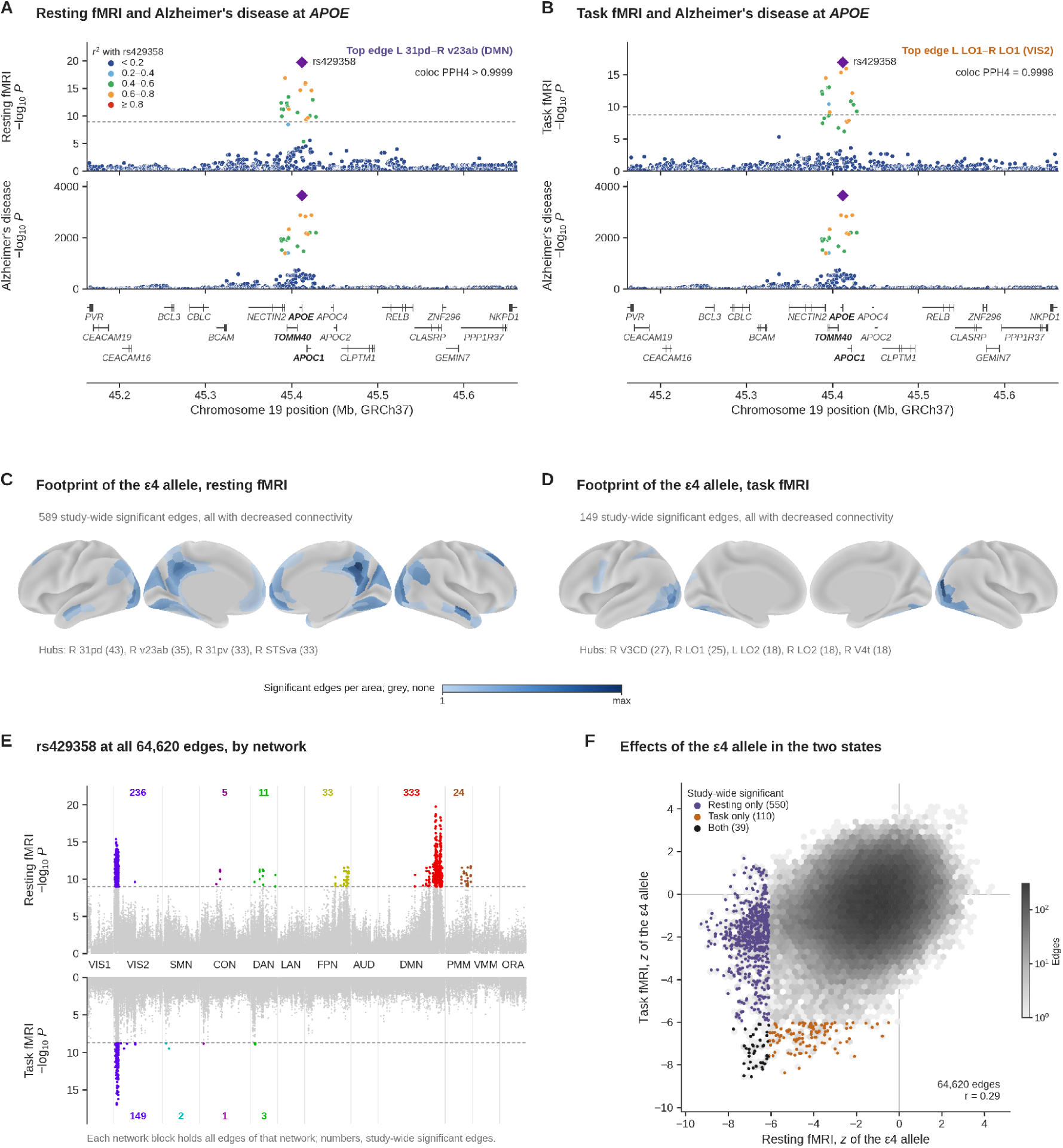
The *APOE* ε4 variant rs429358 is associated with functional connectivity and colocalizes with Alzheimer’s disease. **(A, B)** Regional association plots of the 19q13.32 region for the edge most strongly associated with rs429358 (top) in resting fMRI (**A**; left 31pd–right v23ab, default mode network; *P* = 1.7 × 10^-20^; *n* = 52,983) and task fMRI (**B**; left LO1–right LO1, secondary visual network; *P* = 1.1 × 10^-17^; *n* = 45,010), for the genome-wide association study (GWAS) of Alzheimer’s disease and related dementias (middle; GCST90704647^39^), and for RefSeq genes (bottom). Colors give the linkage disequilibrium (*r*^2^) with rs429358 (diamond). The posterior probability of a shared causal variant (PPH4; coloc.abf) was 0.999998 for resting and 0.9998 for task fMRI, with rs429358 as the shared variant in both. **(C, D)** Footprints of the ε4 allele (C allele of rs429358) in resting (**C**; 589 study-wide significant edges) and task (**D**; 149 edges) fMRI, shown as the number of significant edges per area; the areas with the most edges are listed with their counts. The ε4 allele was associated with lower connectivity at every significant edge in both states. **(E)** Associations of rs429358 with all 64,620 edges in resting (top) and task (bottom) fMRI, grouped by network, so an edge between two networks appears in both blocks. Colored points are study-wide significant edges (dashed lines, threshold), and the numbers give their count for each network. Network abbreviations are as in Fig. 2. **(F)** Effects of the ε4 allele on all edges in resting and task fMRI (*z* scores; Pearson *r* = 0.29). Points mark the edges that were study-wide significant in resting fMRI only (550), in task fMRI only (110), or in both (39). Details are in the Supplementary Note.

Colocalization also linked other loci for disorders and cognition to specific connections (**Fig. 5**). The schizophrenia locus at 2p16.1 (*VRK2*)^40^ colocalized at a left 8Ad–9p default mode connection (PPH4 = 0.905; **Fig. 5A**). An ICA-based study had linked this locus to interactions of the central executive, default mode, and salience networks^22^. A second schizophrenia locus, within *CNTN4* (3p26.3) and new to fMRI GWAS, colocalized at a right PHT–PGs connection (PPH4 = 0.992; **Fig. 5B**). The 2q24.2 locus near *DPP4*, known from earlier fMRI and cognitive GWAS, colocalized at a left PHT–PFm connection with intelligence (PPH4 = 0.982; **Fig. 5C**), educational attainment, cognitive performance, and reaction time (**Table S6**). The bipolar disorder locus in the *FADS* gene cluster (11q12.2)^41^, also new to fMRI GWAS, colocalized at a right TE1p–TE1m connection (PPH4 = 0.970; **Fig. 5D**). A second Alzheimer’s disease locus, near *C16orf95* (16q24.2), also colocalized (PPH4 = 0.972) and was new to fMRI GWAS. Among the 96 task loci, 17 locus-trait pairs at 10 loci colocalized. They included the task 10q26.13 locus near *FAM53B* with educational attainment (PPH4 = 0.938; **Fig. 5E**) and the *EPHA3* locus with cognitive performance (PPH4 = 0.963; **Fig. 5F**).

**Fig. 5.**
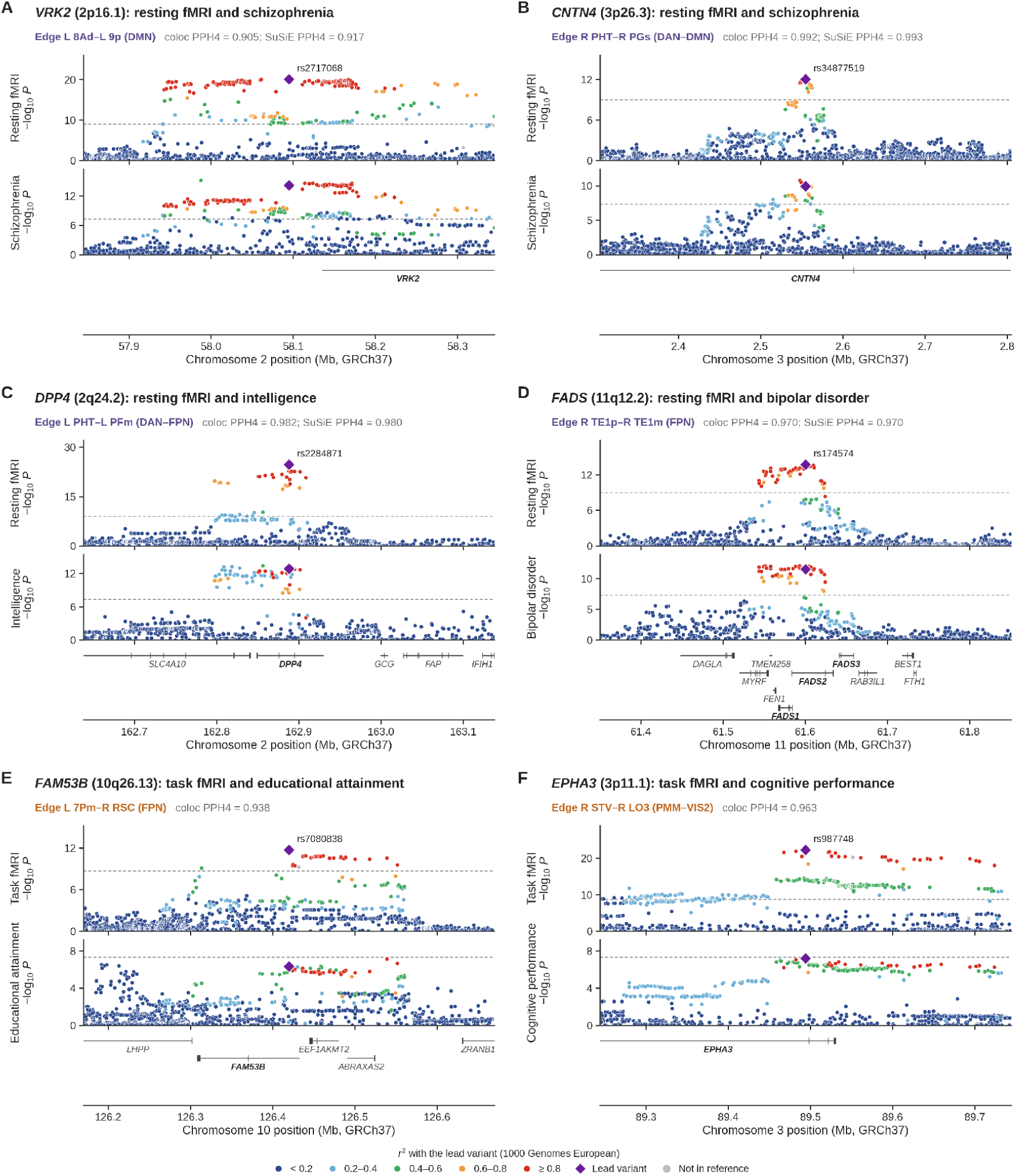
Colocalization of functional connectivity loci with psychiatric and cognitive traits. Regional association plots of six loci with strong colocalization (PPH4 ≥ 0.8 and PPH4 > PPH3) between an edge-level functional connectivity trait (top) and a complex trait (middle), with RefSeq genes (bottom). Each panel shows the edge used in the colocalization analysis, the top edge of the lead variant (Methods). **(A)** *VRK2* (2p16.1): resting fMRI (left 8Ad–left 9p, default mode network) and schizophrenia^72^. **(B)** *CNTN4* (3p26.3): resting fMRI (right PHT–right PGs, between the dorsal attention and default mode networks) and schizophrenia. **(C)** *DPP4* (2q24.2): resting fMRI (left PHT–left PFm, between the dorsal attention and frontoparietal networks) and intelligence^42^. **(D)** *FADS* (11q12.2): resting fMRI (right TE1p–right TE1m, frontoparietal network) and bipolar disorder^41^. **(E)** *FAM53B* (10q26.13): task fMRI (left 7Pm–right RSC, frontoparietal network) and educational attainment^73^. The region also contains *EEF1AKMT2*, the gene assigned to the neighboring resting fMRI locus rs2363340 (Fig. 7F). **(F)** *EPHA3* (3p11.1): task fMRI (right STV–right LO3, between the posterior multimodal and secondary visual networks) and cognitive performance^73^. Edge labels are purple for resting and orange for task fMRI. Above each panel are the PPH4 from coloc.abf (±500 kb; *p*_12_ = 1 × 10^-5^) and, for the resting fMRI loci, the PPH4 from SuSiE-based colocalization (Methods). Variants are colored by their linkage disequilibrium (*r*^2^) with the lead variant (diamond) in the 1000 Genomes European reference panel, and variants absent from the reference panel are shown in gray. Dashed lines indicate the study-wide significance threshold (top; 5 × 10^-8^/*M*_eff_) and *P* = 5 × 10^-8^ (middle). The plots show ±250 kb around the lead variant (GRCh37). Resting fMRI, *n* = 52,983; task fMRI, *n* = 45,010. Details are in the Supplementary Note.

### Genetic correlation maps and circuit closure

Colocalization tests one locus at a time. To map the genome-wide signal of each trait across connections, we estimated the genetic correlation between each edge and 36 complex traits from the LDR summary statistics, with a Bonferroni threshold of *P* < 2.9 × 10^-5^ at rest and 5.0 × 10^-5^ during the task (Methods). In resting fMRI (**Fig. 6A–D**), intelligence was genetically correlated with 934 edges (1.5%), 99% of them positively (**Fig. 6A, C**). Two-thirds of these edges involved the default mode network, mostly connections within it or with the frontoparietal network, the systems for internally directed thought and cognitive control, and the left PHT was their leading hub.

**Fig. 6.**
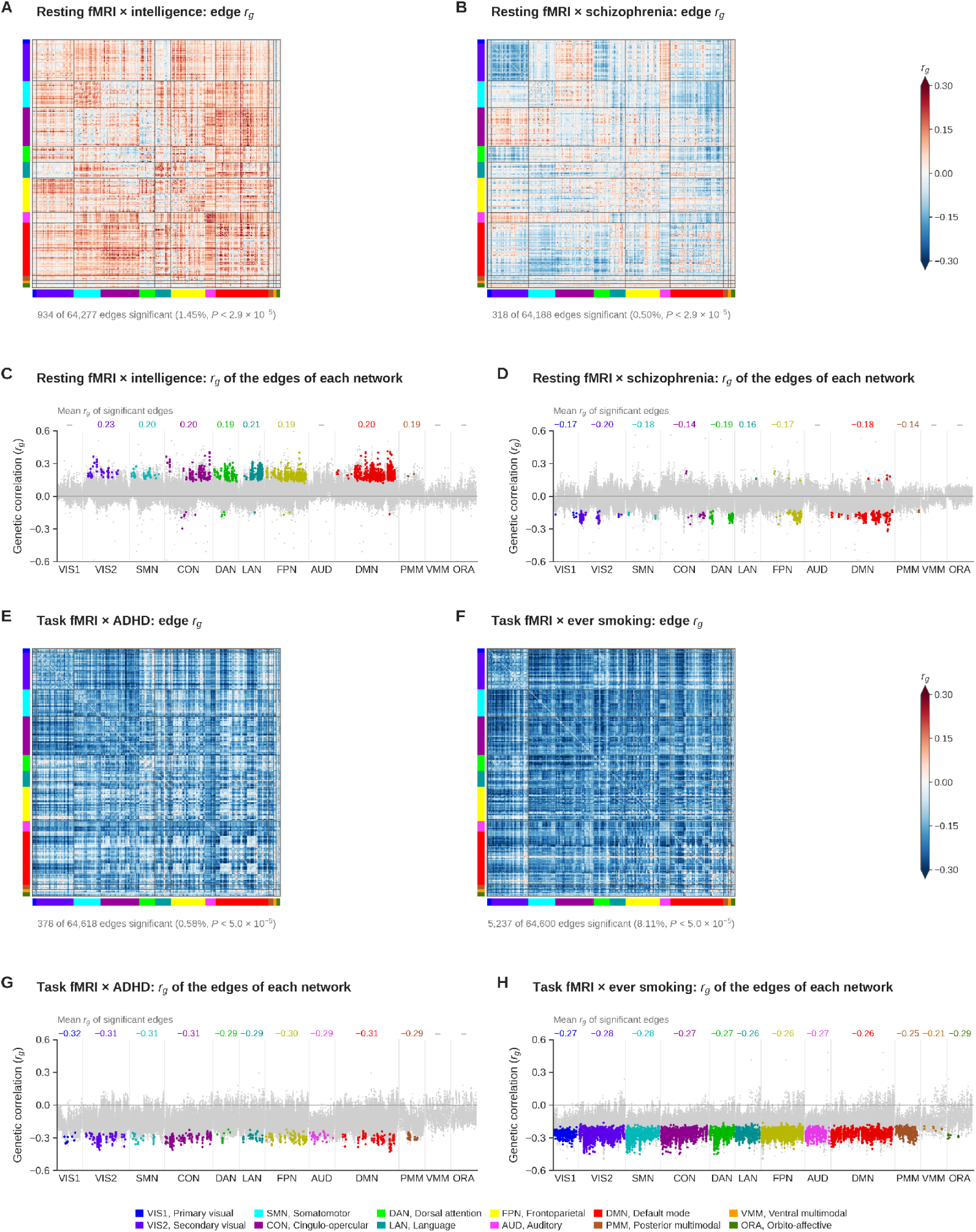
Edge-level genetic correlations of functional connectivity with complex traits. **(A, B)** Genetic correlations (*r*_g_) between resting functional connectivity and intelligence (**A**) and schizophrenia (**B**) for the 64,620 edges, shown as 360 × 360 matrices in which the areas are ordered by network (colored bars). Edges without an estimate and the diagonal are gray, and values beyond ±0.3 are shown in the end colors. The number and percentage of edges with significant genetic correlation (*P* < 2.9 × 10^-5^ at rest and 5.0 × 10^-5^ during the task; Methods) are given below each matrix. **(C, D)** Genetic correlations of the edges of each network with intelligence (**C**) and schizophrenia (**D**) in resting fMRI. Each block contains all edges of one network, so an edge between two networks appears in both blocks. Colored points are significant edges, gray points are nonsignificant edges, and the numbers above the blocks give the mean *r*_g_ of the significant edges (–, no significant edge). **(E, F)** Genetic correlations between task functional connectivity and attention-deficit/hyperactivity disorder (ADHD) (**E**) and ever smoking (**F**). **(G, H)** Genetic correlations of the edges of each network with ADHD (**G**) and ever smoking (**H**) in task fMRI, shown as in **C** and **D**. *M*_eff_ = 46.7 for resting and 26.9 for task fMRI. Network abbreviations are as in Fig. 2. Details are in the Supplementary Note.

Schizophrenia was correlated with 318 edges (0.5%), 97% of them negatively, mainly within the secondary visual network and in default mode connections (**Fig. 6B, D**).

Genetic predispositions to higher intelligence and to schizophrenia were thus associated with higher and lower default mode connectivity at rest, respectively. The two traits share genetic influences across the genome^42^, and these maps place part of that overlap, with opposite signs, on default mode connections. Mendelian randomization with the resting lead variants as instruments gave FDR-significant estimates in the same directions: genetically predicted higher connectivity went with higher educational attainment, cognitive performance, and intelligence, and with lower risk of schizophrenia (**Fig. S1**). Because the instruments were heterogeneous, these estimates serve as directional checks (Supplementary Note). In task fMRI (**Fig. 6E–H**), attention-deficit/hyperactivity disorder (ADHD) was negatively correlated with 378 edges (0.6%; **Fig. 6E, G**) and ever smoking with 5,237 edges (8.1%; **Fig. 6F, H**).

With these maps, we asked whether a colocalized locus acts on the connections that carry the polygenic signal of its trait, which we call circuit closure (Methods). Two loci closed with their traits in both the discovery and the replication samples (**Fig. 7A** and **Table S7**).

**Fig. 7.**
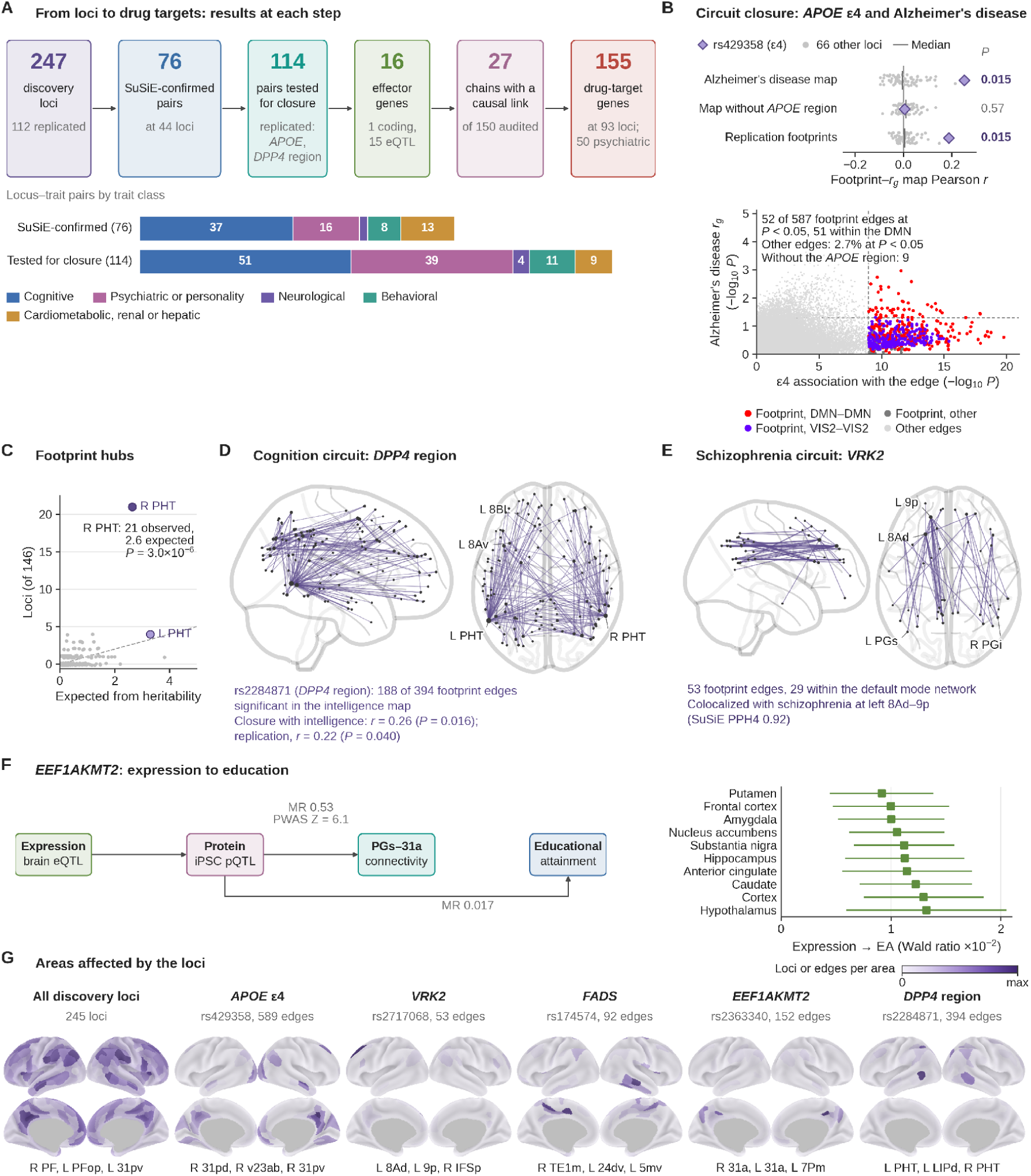
Circuits and genes of the resting-state loci. **(A)** Results at each step (tiles) and trait classes of the SuSiE-confirmed and tested pairs (bars). The last two tiles give the locus-gene-trait chains whose causal link passed, of the 150 audited chains, and the genes with a clinical-stage compound within 500 kb of the discovery loci (**Table 1**). **(B)** Circuit closure of the *APOE* ε4 allele (rs429358) with Alzheimer’s disease. Top, Pearson correlation of its associations across edges (−log_10_ *P*) with the Alzheimer’s disease *r*_g_ map and that of 66 loci without colocalization for the full map, the map without the *APOE* region, and replication footprints; *P*, empirical. Bottom, each edge’s ε4 association against its Alzheimer’s disease *r*_g_ (−log_10_ *P*); footprint edges are colored by network; dashed lines, study-wide significance and *P* = 0.05. **(C)** Principal footprint hubs: discovery loci (of 146 with at least three significant edges) whose footprint centered on each area, against the number expected from heritability (dashed line, equality). **(D)** Footprint edges of rs2284871 (2q24.2, *DPP4* region) that were FDR-significant in the intelligence *r*_g_ map, with its closure (as in B). **(E)** Study-wide significant edges of rs2717068 (2p16.1, *VRK2*). In **D** and **E**, node size increases with the number of edges. **(F)** Expression and protein evidence for *EEF1AKMT2*: Mendelian randomization (MR) estimates of protein abundance on left PGs–31a connectivity and educational attainment and the brain proteome-wide association *Z*-score at that edge (left), and cis-eQTL MR estimates of expression on educational attainment in 10 GTEx brain tissues (right; 95% confidence intervals). **(G)** Areas affected by the loci: discovery loci (245; the two long-range LD proxies of the *EPHA3* locus excluded) whose significant edges included each area (first map) and significant edges per area for the *APOE*, *VRK2*, *FADS*, *EEF1AKMT2*, and *DPP4*-region loci (other maps), each scaled to its maximum. EA, educational attainment. Details are in the Supplementary Note.

The first was the *APOE* ε4 allele, the strongest common genetic risk factor for late-onset Alzheimer’s disease. Its footprint overlapped the Alzheimer’s disease genetic correlation map on the posterior default mode network (Pearson *r* = 0.25, empirical *P* = 0.015; replication, *r* = 0.19, *P* = 0.015; **Fig. 7B**). The shared connections centered on the posterior cingulate cortex (v23ab) and angular gyrus (PGi), the core hubs of the default mode network, and on the anterior superior temporal sulcus (STSva) and lateral temporal cortex (TE1a). These areas support episodic and semantic memory^43,44^, and their connectivity declines in ε4 carriers and across the Alzheimer’s disease spectrum^7,45,46^. Of the 52 footprint edges with a genetic correlation with Alzheimer’s disease, 51 were default mode connections. When the *APOE* region was removed from the Alzheimer’s disease GWAS, the overlap disappeared (*r* = 0.005). The genetic link between Alzheimer’s disease and resting connectivity is therefore carried largely by ε4 and its default mode circuit.

The second was the 2q24.2 locus near *DPP4*, reported in earlier fMRI GWAS, which closed with cognition. Its footprint centered on the left posterior temporal area PHT. The right PHT was a recurrent hub of genetic effects: it was the principal footprint hub for 21 of the 146 loci with at least three significant edges, eight times the number expected from its heritability (*P* = 3 × 10^-6^; **Fig. 7C**), and again the most frequent hub in the replication sample. The hub held against null maps simulated from the principal component basis (Supplementary Note). In the *DPP4*-region footprint, PHT and other dorsal attention areas connected with frontoparietal and default mode areas (**Fig. 7D**).

This temporal, parietal, and frontal circuit overlaps the regions that imaging studies most consistently link to intelligence^47^. The intelligence-increasing allele strengthened 95% of these 394 connections, and 188 of them carry the polygenic signal of intelligence. The locus closed with intelligence (*r* = 0.26, *P* = 0.016; replication, *r* = 0.22, *P* = 0.040; **Fig. 7D**) and cognitive performance (*r* = 0.24, *P* = 0.025), and closure held after the locus was removed from the intelligence GWAS (Supplementary Note). This locus therefore acts on the same circuit as the genome-wide signal of cognition, and its left PHT hub is a priority for functional studies. Among disorders, the *VRK2* schizophrenia locus, also known from earlier fMRI GWAS, acted mainly on default mode connections of the left dorsal prefrontal cortex (areas 8Ad and 9p; **Fig. 7E**).

### From associated loci to genes and candidate drug targets

To connect the loci to genes, we assigned effector genes with a hierarchy of evidence (Methods): a protein-altering variant in the fine-mapped credible set, including pathogenic or risk variants in ClinVar^48^ (Tier 1), or a SuSiE-confirmed colocalization with a brain cis-eQTL in the eQTL Catalogue^49^ (Tier 2). The hierarchy named 16 effector genes (**Fig. 7A** and **Table S8**). With brain cell-type^50^, PsychENCODE^51^, and MetaBrain^52^ expression QTL as further sources, most assignments held, and *FURIN*, a proprotein convertase, was assigned at the bipolar disorder locus 15q26.1 (Supplementary Note).

The genes point to a few biological routes. In single-nucleus expression data^53,54^, *APOE*, the main lipid carrier of the brain, was most highly expressed in astrocytes, and *VRK2*, the long-standing candidate gene at its schizophrenia locus^40^, in oligodendrocytes, which points to glial routes from these loci to connectivity. At the 12q13.12 locus, new to fMRI genetics, the effector gene *RHEBL1* encodes a RHEB-like small GTPase. Higher *RHEBL1* expression predicted higher cognitive performance and intelligence, both in SMR with the HEIDI test and in GTEx brain tissues^55^. Higher *YPEL3* expression likewise predicted less worry. At the major histocompatibility complex (MHC), immune genes carried the signal: the cerebrospinal fluid protein QTL of the butyrophilin *BTN3A2* colocalized with connectivity and with bipolar disorder, schizophrenia, and cognitive performance, and higher BTN3A2 levels and expression went with higher risk of both disorders. In the brain proteome, FHL5 at the *UFL1* locus colocalized with connectivity.

The clearest molecular chain ran through *EEF1AKMT2* at 10q26.13, a locus reported in earlier fMRI GWAS^22^ (**Fig. 7F**). Its expression QTL in the cortex^52^ and its protein QTL in induced pluripotent stem cells^56^ colocalized with the connectivity signal, and the protein QTL also colocalized with educational attainment. Higher protein abundance predicted stronger connectivity between the left angular gyrus (PGs) and posterior cingulate cortex (31a), two hubs of the default mode network (*P* = 2.8 × 10^-20^), and higher educational attainment. Genetically predicted higher expression likewise went with higher educational attainment in all 10 GTEx brain tissues with an instrument^55^ (**Fig. 7F**), and a brain proteome-wide association study showed the same direction at this edge (**Fig. 7F**). EEF1AKMT2 methylates the translation elongation factor eEF1A^57,58^, a core component of protein synthesis. Expression, protein abundance, default mode connectivity, and educational attainment thus rise together, which links the regulation of protein synthesis to this circuit and to cognition (**Fig. 7F**).

To find candidate drug targets, we audited each locus-gene-trait chain for five links: circuit closure, SuSiE-confirmed colocalization, an effector gene or colocalized brain protein QTL, a causal estimate, and a clinical-stage compound in Open Targets^59^. The audit was extended with cell type-specific and PsychENCODE expression QTL^50,51^ and GWAS of migraine, headache, and ischemic stroke (Methods; **Table S9** and **Table S10**). Among the 150 chains of the 247 loci, the causal link passed for 27 (**Fig. 7A**), including *RHEBL1* with cognition, *GTDC1* with cognitive performance, and *YPEL3* with worry. The cortical areas affected by all discovery loci and by the *APOE*, *VRK2*, *FADS*, *EEF1AKMT2*, and *DPP4*-region loci are mapped in **Fig. 7G**.

Three genes stood out for brain disorders. *APOE* was supported for Alzheimer’s disease by colocalization at rs429358, the protein-altering ε4 isoform, colocalization with the ApoE protein QTL in cerebrospinal fluid (PPH4 = 0.9998), and circuit closure. It also has a direct therapeutic link: *APOE2* gene therapy^60^ has completed a phase 1/2 trial in ε4 homozygotes (LX1001; NCT03634007), and anti-amyloid antibodies act on the amyloid pathway downstream of *APOE*^61,62^ (**Table 1**). At the *FADS* locus, new to fMRI genetics and colocalized with bipolar disorder^41^, the variant shared with connectivity raised *FADS1* expression in oligodendrocytes and cortex^52^ and lowered disease risk. The same allele weakened connectivity in the cingulo-opercular and somatomotor networks and shifted plasma fatty acids toward the products of desaturation, arachidonic and omega-3 fatty acids (Supplementary Note). This chain links polyunsaturated fatty acid synthesis to connectivity and to bipolar disorder. *FURIN*, a proprotein convertase at 15q26.1, was also supported for bipolar disorder, and its higher expression predicted lower risk. *FADS1* and *FURIN* are tractable drug targets^59^, and their protective direction of effect points to activation. *RHEBL1* is also tractable, and its higher expression went with higher cognitive scores and lower ADHD risk.

Beyond these genes, 155 genes within 500 kb of the loci are targets of clinical-stage compounds (**Table 1**). Compounds of 50 of them had been tested for psychiatric disorders, and 15 were targets of approved psychiatric drugs, including *SLC6A4* (antidepressants), *HTR2A* (antipsychotics), and *CACNA2D2* and *CACNB3* (pregabalin for anxiety). For seven genes, the compounds had been tested for the trait that colocalized with the locus. At the 10q25.3 locus, new to fMRI genetics and colocalized with ADHD, *SLC18A2* is the target of amphetamines approved for ADHD. At an MHC locus colocalized with major depressive disorder, TNF inhibitors (infliximab and golimumab) have been tested in depression. For nine genes at cognition loci, compounds had been tested for cognitive impairment, including the DPP4 inhibitor sitagliptin at 2q24.2. Brain expression data also tied three drug targets to their loci: *CRHR1* and *MAPT* at 17q21.31, colocalized with educational attainment, and *MAPKAPK5* at 12q24.12, colocalized with reaction time.

### Comparison of resting-state and task-state connectivity

To see how brain state shapes genetic effects, we compared resting and task fMRI edge by edge (**Fig. 8A** and **Fig. S2**). Task connectivity came from a shorter scan and, because it was computed over all conditions of a block-design task, combines background connectivity with stimulus-evoked coactivation^16,63^. Differences between the states therefore reflect measurement as well as brain state. Nearly all edges were heritable in both states, but resting heritability was higher: 39.9% of resting and 3.8% of task edges had *h*^2^ ≥ 10% (**Fig. S2A, B**), and resting heritability exceeded task heritability at 66.9% of the edges. The two heritability maps were weakly correlated (*r* = 0.10; **Fig. 8A**). Averaged over the connections of each network, resting heritability was higher in all 12 networks, most markedly in association networks (**Fig. S2C**). For connections within the somatomotor and auditory networks, task heritability was higher (7.3% versus 4.4% and 8.2% versus 2.7%; **Fig. S2D**).

**Fig. 8.**
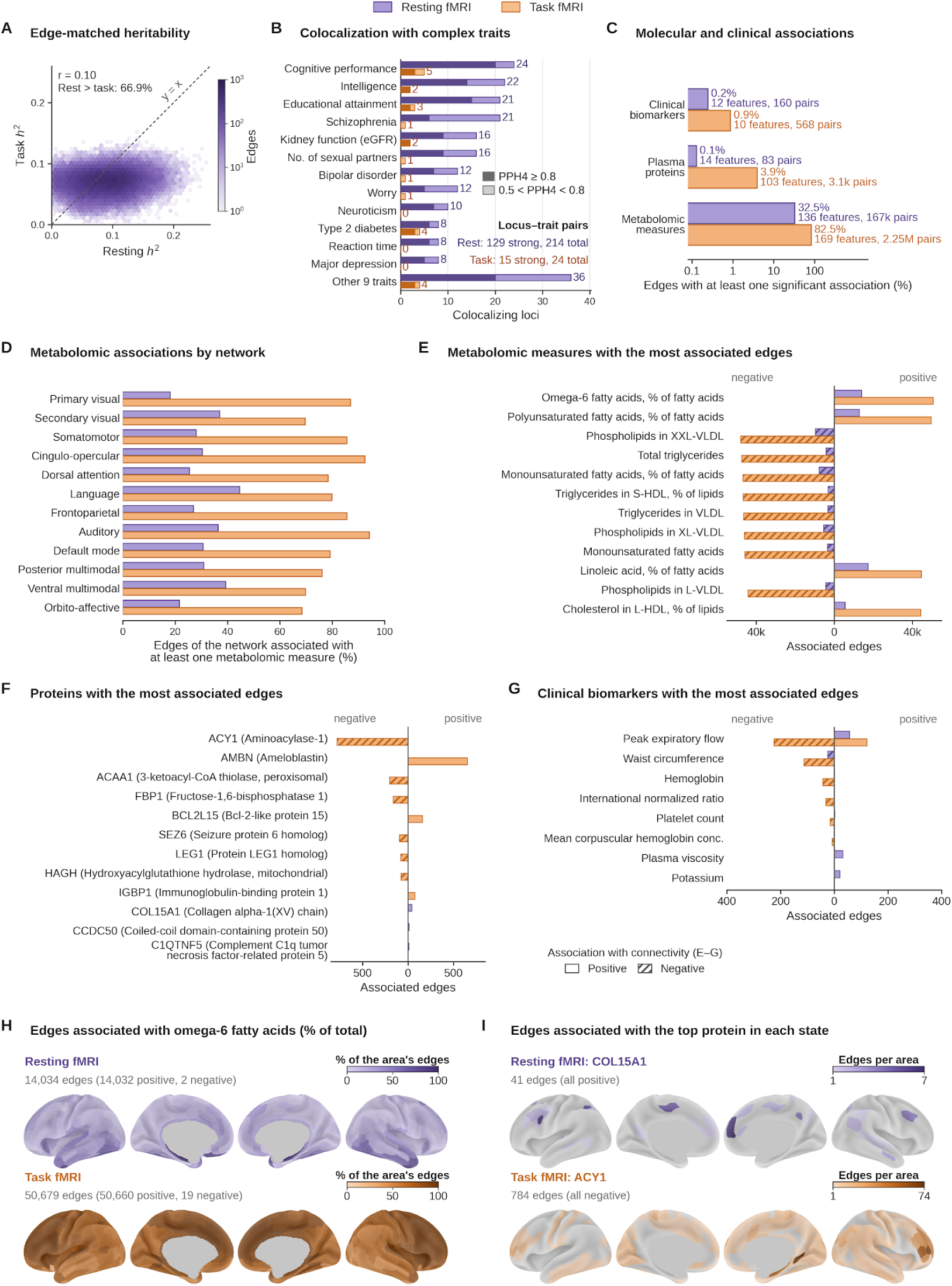
Heritability, colocalization, and molecular and clinical associations of resting and task functional connectivity. **(A)** SNP heritability of the 61,930 edges with estimates in both states (resting fMRI, *n* = 52,983; task fMRI, *n* = 45,010). Hexagons are shaded by edge count (log scale); dashed line, equal heritability; Rest > task, percentage of edges more heritable at rest. **(B)** Numbers of loci colocalizing with each complex trait^35^ at PPH4 ≥ 0.8 and PPH4 > PPH3 (dark segments) or 0.5 < PPH4 < 0.8 (light segments), among all 247 resting and 96 task loci, for the 21 traits tested at all loci. The 12 traits with the most colocalizing resting loci are shown individually, and the other 9 are pooled (locus-trait pairs). Inset, locus-trait pairs (total, PPH4 > 0.5). **(C)** Percentage of the 64,620 edges with at least one significant association (*P* < 0.05/(*M*_eff_ × *K*); *K*, features per layer) with clinical biomarkers, plasma proteins, or metabolomic measures, with the numbers of associated features and edge-feature pairs (log scale). **(D)** Percentage of the edges of each network associated with at least one metabolomic measure. **(E–G)** Edges with positive (unhatched) and negative (hatched) associations at rest (purple) and during the task (orange) for the metabolomic measures (**E**) and proteins (**F**) with the most associated edges, and for clinical biomarkers with at least 8 associated edges (**G**). **(H)** Percentage of the 359 edges of each area associated with omega-6 fatty acids (percentage of total fatty acids), on a common scale. **(I)** Number of associated edges per area for the protein with the most associated edges in resting (COL15A1) and task (ACY1) fMRI, scaled separately. In **H** and **I**, gray marks the medial wall and areas without associated edges. Details are in the Supplementary Note.

Genetic discovery followed the same pattern (247 resting and 96 task loci; **Fig. S2E**). Of the 96 task loci, 28 lay within 500 kb of a resting locus, five with identical lead variants. Shared variants affected far fewer connections during the task: rs429358 (*APOE*) affected 589 resting and 149 task edges, and the *EPHA3* variants affected 2,867 to 2,889 resting and 257 to 279 task edges (**Fig. S2F**). During the task, most of these footprints shifted toward the visual cortex. The emotion-processing task engages this cortex strongly, so stimulus-evoked coactivation is expected there.

Links to complex traits also differed between the states. Resting connectivity had more significant genetic correlations than task connectivity for schizophrenia (0.50% of edges versus none) and intelligence (1.45% versus 0.07%), whereas task connectivity had more for ADHD (0.58% versus 0.02%; **Fig. S2G, H**). Mendelian randomization gave FDR-significant estimates for 7 of 14 outcomes with the resting loci and none with the task loci (**Fig. S1** and **Table S11**). Across the 21 traits tested at all loci (Methods), 129 locus-trait pairs at 65 of the 247 resting loci colocalized strongly, compared with 15 pairs at 9 of the 96 task loci (**Fig. 8B**; task loci in **Table S12**). Resting-state connectivity is thus more heritable than task-state connectivity, yields more loci, and colocalizes more often with complex traits.

Phenotypic associations with circulating molecular traits showed the opposite pattern. We tested each edge for association with 49 clinical biomarkers, 2,922 plasma proteins, and 251 nuclear magnetic resonance (NMR) metabolomic measures in the UKB. The Bonferroni threshold was *P* < 0.05/(*M*_eff_ × *K*), where *K* is the number of features in the layer (Methods). Task connectivity had 37.1-fold more protein-edge, 13.4-fold more metabolite-edge, and 3.6-fold more clinical biomarker-edge pairs than resting connectivity (**Fig. 8C, D** and **Table S13**). Fatty acid composition and triglyceride-rich lipoproteins led the metabolomic associations (**Fig. 8E**). The protein associations differed between the states (**Fig. 8F**), and peak expiratory flow led the biomarkers in both (**Fig. 8G**). During the task, the metabolite associations extended to most of the connectome; omega-6 fatty acids, for example, were associated with 14,034 resting and 50,679 task edges (**Fig. 8H** and **Table S14**). The edges of the leading protein in each state, COL15A1 at rest and ACY1 during the task, are mapped in **Fig. 8I**.

The metabolite associations agreed between the states connection by connection. Of the resting edges with a metabolite association, 83% also had one during the task, and for each leading fatty acid measure, the direction agreed at 99.8% to 100% of the shared edges. Each measure had the same sign at nearly all of its edges (at least 95% of them for 167 of the 169 task measures). The associations shared by the two states were stronger during the task (median |*Z*|, 7.2 versus 5.2), even with a smaller sample (median *n*, 22,449 versus 26,236). Associations of one sign across most of the connectome fit a whole-brain component of connectivity that reflects systemic physiology, including the vascular contribution to BOLD coupling^64,65^. In this account, the coactivation evoked by a block-design task amplifies the component. Task-state connectivity is thus less heritable than resting-state connectivity but more closely coupled to circulating metabolic traits, in line with a larger state-dependent component of connectivity during a task^66^.

## DISCUSSION

We present a connection-level genetic atlas of the human cerebral cortex in two brain states. The atlas comprises summary statistics for all 64,620 connections, 247 resting loci (193 new to fMRI genetics) and 96 task loci with their footprints, and edge-level genetic correlation maps for 36 traits. Three findings stand out. First, genetic effects on connectivity are largely connection-specific: most loci affected few edges, and most acted on connections between networks. The coupling between networks is itself heritable, and traits that aggregate connections average out this signal. Second, genetic predispositions to higher intelligence and to schizophrenia are associated with higher and lower default mode connectivity, respectively, and the 2q24.2 locus near *DPP4* acts on the connections that carry the polygenic signal of intelligence. Its footprint centers on the left posterior temporal area PHT, and the right PHT is a recurrent hub of genetic effects; both are priorities for functional studies. Third, the *APOE* ε4 allele lowers connectivity across the posterior default mode and secondary visual networks, and this circuit effect persists after adjustment for mean connectivity. Its direction matches the connectivity changes reported in ε4 carriers and across the Alzheimer’s disease spectrum^7,45,46^. The default mode connections of this footprint carry the genetic correlation of Alzheimer’s disease with connectivity and are candidate imaging phenotypes for genetic and clinical studies.

Circuit closure, which underlies the second and third findings, revealed two kinds of link between a locus and the polygenic signal of its trait. At *APOE*, one locus carries the genetic correlation between Alzheimer’s disease and connectivity: removing the *APOE* region from the disease GWAS abolished the genetic correlation at the footprint connections. Near *DPP4*, the 2q24.2 locus acts on the same connections as the polygenic signal of intelligence, in both samples and independently of its own contribution to the map. The first kind places a known genetic correlation on specific connections; the second points to circuits shared by a locus and the polygenic signal of its trait.

At the molecular level, the loci point to candidate mechanisms. For *EEF1AKMT2*, at a locus known from earlier fMRI GWAS^22^, higher expression and protein abundance (measured in induced pluripotent stem cells) were linked to higher connectivity and educational attainment. This sequence implicates the regulation of translation elongation. *FADS1*, at a bipolar disorder locus new to fMRI GWAS, links fatty acid desaturation to cingulo-opercular and somatomotor connectivity and to disease risk, in agreement with Mendelian randomization linking arachidonic acid synthesis to bipolar disorder^67^.

The drug-target analysis links the loci to existing compounds and to new candidates. Within 500 kb of the loci, 155 genes are targets of clinical-stage compounds (**Table 1**). For seven of them, compounds had been tested for the trait that colocalized with the locus: amphetamines, which act on *SLC18A2*, are approved for ADHD, and TNF inhibitors have been tested in major depression. These links are positional and mark compounds for repurposing studies that first confirm the gene. For three drug targets, *CRHR1* and *MAPT* at 17q21.31 and *MAPKAPK5* at 12q24.12, brain expression data also tied the gene to the locus. Among the effector genes, *APOE* has a direct therapeutic link through *APOE2* gene therapy. Tractable genes with genetic support point to new targets: higher *RHEBL1* expression went with higher cognitive scores and lower ADHD risk, and at two bipolar disorder loci the protective direction of effect of *FADS1* and *FURIN* calls for activation^68^. A complete chain from circuit to drug will need compounds for these genes and gene-level evidence for the existing drug targets.

These findings come mainly from resting-state connectivity, which shares a group-level architecture with task-state connectivity^69,70^ but differs from it genetically. The same variants affected fewer connections during the task, mainly in the visual cortex that the task engages^18^. With its higher heritability and more frequent disease colocalization, resting connectivity is the better substrate for genetic discovery. Task connectivity, in turn, had stronger and more widespread associations with circulating metabolites, and each metabolite’s associations had the same sign across most of the connectome. This pattern points to a whole-brain, state-dependent component of connectivity that reflects systemic physiology, which makes task connectivity informative for studies of physiological state. Jointly modeling the two states may separate stable from state-dependent genetic influences.

The LDR framework made the edge-level analysis feasible and localized genetic effects to the connections that whole-brain ICA^20–22^ and aggregated parcellation traits combine.

For example, the 2q13 locus near *PAX8*, also detected with ICA^22^, affected 24,888 edges, whereas the *VRK2* locus affected 53 edges, mostly in the default mode network.

Researchers can use the atlas to place loci and traits on specific connections and to choose imaging endpoints. As samples grow, circuit closure and target chains can be tested for new loci with independent footprints and leave-locus-out maps.

Several considerations frame these results. Edge-level effects are reconstructed from 300 principal components per part, which explained 78% to 80% of the edge variance in each part. The basis retains a median of 88% of an effect that spans all edges of an area. Of an effect confined to a single edge, it retains about 2%, and 53% to 70% of the remainder falls on edges that share an area with that edge. Footprints therefore resolve genetic effects at the level of areas and their connections (Supplementary Note). The analyses used one parcellation and the autosomes, cell-type enrichment used annotations of four adult cortical cell types, and colocalization was tested at the top edge of each locus. The replication sample reproduced the sign of 99.6% of the study-wide significant effects, and in circuit closure, independent footprints and leave-locus-out maps controlled for the contribution of a locus to the map it was compared with. The task loci await replication in an independent task fMRI sample. The task data came from a single emotion-processing task^18,71^, and task connectivity includes stimulus-evoked coactivation and is less reproducible and heritable than resting connectivity^20,63^. Participants were middle-aged to older adults of European ancestry, and UK Biobank volunteers are healthier than the general population, and the associations with circulating molecular traits describe phenotypic co-variation in a cross-sectional sample. Larger, more diverse datasets with multiple tasks will show whether these results generalize across ages and ancestries.

In summary, the atlas created here moves the genetics of cortical functional connectivity from aggregated traits to individual connections. It places loci for cognition and disease on specific circuits, distinguishes the stable genetic architecture of resting connectivity from the state-dependent component of task connectivity, and provides summary statistics with which these circuits can be tested in new samples.

Note: The Supplementary Information (one PDF file with the Supplementary Note and Supplementary Figures S1–S22) and the Supplementary Tables (one Excel workbook with Tables S1–S25) are available.

## Data Availability

Access to the individual-level UK Biobank data used in this study is controlled by UK Biobank and is subject to its eligibility requirements, approval procedures, and data-use conditions. Information about data access is available at https://www.ukbiobank.ac.uk/use-our-data/apply-for-access/. The authors do not redistribute individual-level participant data. Study results are reported in the manuscript and accompanying Supplementary Information and Supplementary Tables.

## ACKNOWLEDGEMENTS

We thank the research assistants in the UNC BIG-S2 team for downloading and preprocessing raw images. We thank the UK Biobank participants and the research teams for collecting, processing, and sharing the data. We thank the University of North Carolina at Chapel Hill and its Research Computing groups for computational resources and support. This research has been conducted using the UK Biobank Resource under Application Number 22783.

This work was partially supported by the National Institutes of Health (NIH) grants R01AG085581 and RF1AG082938 (T.L., B.Z., and H.Z.), U01AG088667 (T.L., H.Z., and J.S.), RF1AG098697 (T.L. and H.Z.), R01MH136055 (H.Z., B.Z., and T.L.), K01AG095286 and R21HD120911 (T.L.), and U01HG011720, R01MH125236, and P50HD103573 (Y.L.). The content is solely the responsibility of the authors and does not necessarily represent the official views of the NIH.

The icons in Fig. 1 are from Bioicons (bioicons.com): DNA double helix by James Lloyd (CC0); GWAS plot and LD block by Ana Viñuela (CC0); database from OpenClipart (CC0); pill and blood sample tube by Marcel Tisch (CC0); dimensionality reduction and heatmap by Chenxin Li (CC BY 4.0); scatter plot by dbs-sticky (MIT); and brain, chromosome, and protein from Servier Medical Art (CC BY 3.0).

## AUTHOR CONTRIBUTIONS

H.Z., B.Z., and T.L. conceived the study. S.W. performed the data analyses, with contributions from T.L., Z.J., A.H., Z.S., X.L., S.G., S.L., S.Y., X.Q., I.P., Y.C., Y.J., B.T., Z.Z., S.H., Y.Z., P.G., M.H., and Y.D. contributed to UKB imaging data processing. S.W. and B.Z. wrote the manuscript. H.Z., B.Z., T.L., Y.L., J.S., and P.S. critically revised the manuscript.

## METHODS

### Imaging data

We used the resting and task fMRI data of the UKB, which obtained ethics approval from the North West Multicentre Research Ethics Committee (approval number: 11/NW/0382), and all participants provided written informed consent. Image acquisition and preprocessing are described in the Supplementary Note. The task fMRI used the emotion-processing task^71,74^, and connectivity was computed over all task conditions. The preprocessed time series on the cortical surface were parcellated into the 360 areas of the Glasser atlas^1^. The functional connectivity between each pair of areas was computed with FSLNets^75^ as the full correlation of their time series, converted by Fisher’s r-to-z transformation and scaled as in the UKB imaging pipeline^76^. The 64,620 upper-triangular edges were analyzed as traits, and the areas were grouped into 12 functional networks^30^ (Table S1).

### Samples

The discovery samples were drawn from the 56,614 and 47,976 UKB participants with resting and task fMRI from imaging phases 1 to 6. We included participants of self-reported white ethnic background (hereafter European ancestry) and excluded those flagged by the UKB as outliers for heterozygosity or missing rate, with sex-chromosome aneuploidy, or with discordant genetic and reported sex. After removal of participants with missing data, 53,096 (resting) and 45,115 (task) participants remained, of whom 52,983 and 45,010 were included in the association analyses, depending on the part of the connectome. The replication sample comprised 15,546 white British participants with resting fMRI from imaging phase 7, none of whom were in the discovery sample (Supplementary Note).

### Edge-level genome-wide association analysis

Association analyses were performed with HEIG^28^, separately for resting and task fMRI and for four parts of 16,155 edges. Principal component analysis (PCA) of the mean-centered edges of each part in unrelated participants gave the bases (eigenvectors), onto which the connectivity of each participant was projected to construct LDRs, adjusting for age, age^2^, sex, their interactions, the top 40 genetic principal components, and assessment centre. This step used the functional PCA module of HEIG with kernel smoothing disabled, which is equivalent to standard PCA. The top 300 LDRs of each part were tested for association with 7,847,434 imputed autosomal variants (UKB imputation version 3; GRCh37) by linear regression with leave-one-chromosome-out adjustment for relatedness. After removal of non-SNP and strand-ambiguous variants, 6,633,888 SNPs remained, and the association of each SNP with each edge was reconstructed from the LDR summary statistics. The effective number of independent edges was computed from the eigenvalues *λ* of each part as *M*_eff_ = (Σ*λ*)^2^/Σ*λ*^229^ and summed over the parts (46.7 for resting and 26.9 for task fMRI). Study-wide significance was defined as *P* < 5 × 10^-8^/*M*_eff_. Software versions and further details are given in the Supplementary Note.

Imputed variants were retained when the minor allele frequency was ≥ 0.01, the imputation INFO score ≥ 0.8, variant missingness ≤ 10% and Hardy–Weinberg equilibrium *P* ≥ 1 × 10^-7^, and participants were retained when genotype missingness was ≤ 10%.

### Definition of loci and replication

Study-wide significant variants were retained when the number of edges with *P* < 1 × 10^-5^ exceeded a null quantile estimated by HEIG. They were clumped with PLINK^77^ using the 1000 Genomes European reference panel (lead variants at *r*^2^ < 0.1 within 500 kb), within and across the four parts. Lead variants within 500 kb of each other were merged into one locus. The top edge of a lead variant was the edge with its smallest *P* value within the part in which the locus was identified. Its footprint comprised its study-wide significant edges, and circuit closure used its associations with the edges that had genetic correlation and heritability estimates (61,933 to 61,936 of the 64,620 edges). In the replication sample, connectivity was projected onto the discovery bases and analyzed in the same way. A locus replicated at the nominal level when its lead variant was associated with its top edge at *P* < 0.05 with the same direction of effect, and at the Bonferroni level when *P* < 0.05/247. Long-range LD proxies and the conditional analysis of the 17q21.31 region with GCTA-COJO^78^ are described in the Supplementary Note.

### Comparison with earlier GWAS

The lead variants were compared with the genome-wide significant associations (*P* ≤ 5 × 10^-8^) in the GWAS Catalog (release 2026-08-24) and with the loci reported in earlier fMRI GWAS^20–22,24–26^, counting associations listed in supplementary tables when they reached the significance threshold of their own study. A locus was considered previously reported when an fMRI association lay within 500 kb of its lead variant (Supplementary Note).

### Heritability and genetic correlation

Edge-level SNP heritability and genetic correlations with 36 complex traits (Table S15) were estimated with HEIG from the LDR summary statistics, using LD matrices of 1,159,563 HapMap3 variants estimated in 50,000 unrelated UKB participants of European ancestry. Heritability was tested at an FDR of 5% across edges. Genetic correlations allowed for sample overlap, and significance was defined as *P* < 0.05/*M*_eff_/37 for all traits, a Bonferroni correction for the 37 GWAS tested, of which liver disease had no valid SNP-heritability estimate (Supplementary Note).

### Functional enrichment

Enrichment analyses used the genome-wide summary statistics of one edge per locus, the edge with the most study-wide significant variants within 250 kb of the lead variant (228 distinct edges for the resting and 95 for the task loci).

Because LD score regression is linear in the χ^2^ statistic, each state was analyzed as the mean χ^2^ across its edges, with block-jackknife standard errors. Stratified LD score regression^32^ with the baselineLD v2.2 model^79^ tested the ATAC, H3K27ac, and H3K4me3 peaks of neurons, astrocytes, oligodendrocytes, and microglia^33^, each against the union of the four, and estimated the fold enrichment of the union in a model with the baselineLD annotations. It also tested the specifically expressed genes of 54 GTEx tissues, each against all expressed genes^55,80^. MAGMA^34^ gene statistics were combined across edges (Table S16) and tested against 10,626 MSigDB gene sets^81^. Details are given in the Supplementary Note.

### Colocalization

Colocalization between connectivity and 23 complex traits and diseases (Table S17) was tested with coloc.abf^35^ (*p*_1_ = *p*_2_ = 1 × 10^-4^ and *p*_12_ = 1 × 10^-5^) in windows of ±500 kb around each lead variant, at the top edge of the lead variant. Pairs were tested when at least 50 variants were shared and the external GWAS had a regional association (*P* < 1 × 10^-4^), and strong colocalization was defined as PPH4 ≥ 0.8 and PPH4 > PPH3.

Strong pairs of the resting loci were refitted with SuSiE-based colocalization^36^ (*L* = 5), with LD from the 1000 Genomes European reference panel^82^. For Alzheimer’s disease, we used the consensus meta-analysis of Alzheimer’s disease and related dementias without proxy cases^39^. Coronary atherosclerosis and liver disease were tested at the 112 replicated resting loci and the task loci, and the other 21 traits at all loci. Sensitivity analyses of the prior and of the tested edge are described in the Supplementary Note.

### Multi-omics association analysis

Edge-level associations with 49 clinical biomarkers, 2,922 plasma proteins (Olink), and 251 NMR metabolomic measures were estimated with the LDR framework in unrelated participants of European ancestry with imaging and omics data, adjusting for the covariates of the association analyses. Significance was defined as *P* < 0.05/(*M*_eff_ × *K*), where *K* is the number of features in the layer, and features measured in fewer than 1,000 participants were excluded (Supplementary Note).

### Mendelian randomization

Two-sample Mendelian randomization was performed with TwoSampleMR^83^, using the lead variants of the resting or task loci as instruments and GWAS of 14 cognitive, psychiatric, neurological, and sleep-related traits as outcomes (Table S18). The association of each instrument with connectivity was taken from the edge with the most study-wide significant variants near its lead variant. We report inverse-variance weighted (fixed-effect and multiplicative random-effects), MR-Egger, weighted median, and weighted mode estimates, with FDR correction across the 14 outcomes in each state (Supplementary Note).

### Circuit closure

For each locus-trait pair with strong colocalization and a usable edge-level genetic correlation map (114 pairs at 59 loci for 18 traits), the footprint of the lead variant (−log_10_(*P*) at each edge) was compared with the genetic correlation map of the trait (−log_10_(*P*)) by the Pearson correlation over the edges with estimates. Null models that permuted the areas within each of the 12 functional networks^30^ or rotated the area centroids on the sphere and reassigned them one to one^84–86^ were too liberal, because footprints share edge-level structure with the maps (Supplementary Note).

Each pair was therefore compared with all other discovery loci that did not colocalize with the trait (PPH4 < 0.5; outside the extended MHC region, chromosome 6 at 25–34 Mb, GRCh37; the two long-range LD proxies of the *EPHA3* locus excluded), giving an empirical *P* value and a *z* score. *P* values were adjusted with the Benjamini–Hochberg procedure across all pairs. For each trait with at least three colocalized loci (PPH4 ≥ 0.8, outside the MHC), the closure of the colocalized loci was compared with that of the other loci by a one-sided Mann–Whitney test, with Benjamini–Hochberg adjustment across traits. Sensitivity analyses used SuSiE-confirmed loci only or partialled edge SNP heritability out of both maps. The pair and class tests were repeated with footprints re-estimated in the replication sample from its LDR summary statistics with the discovery bases; hubs in that sample used edges at *P* < 10^-5^. For the *APOE*–Alzheimer’s disease pair and 15 other pairs (Table S7), the genetic correlation maps were re-estimated after removing the variants within 1 Mb of the lead variant from the trait GWAS. At the *APOE* locus, footprint edges with nominally significant genetic correlations with Alzheimer’s disease (*P* < 0.05) were compared with the other edges by Fisher’s exact test. The principal footprint hub of a locus was the area with the most study-wide significant edges; the number of loci per hub was compared with that expected from nodal SNP heritability (mean heritability of the incident edges) by a Poisson model with Pearson overdispersion (Supplementary Note).

### Effector genes

At the 66 loci with at least one strong colocalization, the connectivity signal was fine-mapped with SuSiE, and the credible-set variants were annotated with Ensembl VEP^87^. A gene was placed in Tier 1 if it carried a protein-altering credible-set variant (posterior inclusion probability ≥ 0.1 in both the connectivity and the trait fits; predicted deleterious, or a missense variant classified in ClinVar^48^ as pathogenic, likely pathogenic, or a risk factor). It was placed in Tier 2 if its expression QTL in any of 20 brain datasets of the eQTL Catalogue^49^ colocalized with connectivity (coloc.abf screen and SuSiE confirmation). A gene was assigned when it was the only gene in the highest tier at its locus; ties were resolved by protein-coding biotype and then by a margin of at least 0.10 in the expression QTL PPH4. The assignment was repeated in five settings of fine-mapping and expression QTL screening, and with expression QTL of eight brain cell types, PsychENCODE, and MetaBrain as additional Tier 2 sources (Supplementary Note).

### Expression- and protein-level causal analyses

Cis-eQTL Mendelian randomization was performed for the 15 Tier 2 effector genes in the 13 brain tissues of GTEx v8^55^, with Steiger-filtered instruments^88^ and Benjamini–Hochberg adjustment across all estimates. For each Tier 2 gene, SMR also used the top cis-eQTL (*P* < 5 × 10^-8^) in each of the 20 brain datasets of the eQTL Catalogue; a dataset supported the causal link when the Benjamini–Hochberg-adjusted SMR *P* value across datasets was at most 0.05 and the HEIDI *P* value was at least 0.05. Protein QTL Mendelian randomization and colocalization used the HipSci induced pluripotent stem cell proteome^56^, cerebrospinal fluid ApoE and BTN3A2^89^, plasma DPP4^90^, the UKB Pharma Proteomics Project^91^, and the dorsolateral prefrontal cortex proteome^92^, which was tested for all protein-coding genes within 500 kb of the loci with strong colocalization. A brain proteome-wide association study used FUSION weights^92,93^ (Table S19; Supplementary Note), and the protein QTL of each association was tested for colocalization with the edge signal (coloc.abf and SuSiE).

Colocalization with MetaBrain eQTL of five brain tissues^52^ used SuSiE on variants whose meta-analysis sample size was at least 90% of the gene maximum.

### Target-chain audit

Each chain linked a locus, a candidate gene, and a colocalized trait (150 chains). The circuit link required closure above that of the other discovery loci (empirical *P* < 0.05); the disease link, SuSiE-confirmed colocalization; the gene link, a Tier 1 or Tier 2 gene or a brain protein QTL that colocalized with the edge; the causal link, a significant and directionally consistent cis-eQTL or protein QTL Mendelian randomization estimate; and the drug link, a drug or clinical candidate for the gene in the Open Targets Platform (release 26.09)^59^. A chain was complete when all five links passed and the estimated directions of effect agreed. The audit was then expanded with the 2,204 protein-coding genes within 500 kb of the loci, cell type-specific and PsychENCODE expression QTL^50,51^, and GWAS of migraine, headache, stroke, and ischemic stroke. The re-scored audit comprised 295 chains (Supplementary Note). Clinical-stage compounds were matched to the colocalized trait by their Open Targets indications. Psychiatric indications were schizophrenia or psychosis, bipolar disorder, depression, anxiety, ADHD, autism, obsessive-compulsive or post-traumatic stress disorder, substance use, eating disorders, insomnia, Tourette syndrome, and agitation or delirium, each with its highest clinical stage in Open Targets.

### Display items

Display details of the main figures, including color scales, exclusions, and tie-breaking rules, are given in the Supplementary Note.

## Code availability

The HEIG software^28^ is available at https://github.com/Zhiwen-Owen-Jiang/heig.

## Data availability

Individual-level UK Biobank data are available to approved researchers through the UK Biobank. The GWAS summary statistics generated in this study will be submitted to the NHGRI-EBI GWAS Catalog: the edge-level summary statistics for all 64,620 edges in both states, and the LDR summary statistics and bases from which they are reconstructed.

Footprints of all loci, edge-level SNP heritability and edge-level genetic correlation maps for 36 traits will be deposited in Zenodo on publication. External GWAS summary statistics are listed in Tables S15, S17, and S18, and the QTL datasets and the GWAS added in the expanded audit are described in the Supplementary Note.

